# Artificial Intelligence for Advance Requesting of Immunohistochemistry in Diagnostically Uncertain Prostate Biopsies

**DOI:** 10.1101/2021.02.20.21252126

**Authors:** Andrea Chatrian, Richard T Colling, Lisa Browning, Nasullah Khalid Alham, Korsuk Sirinukunwattana, Stefano Malacrino, Maryam Haghighat, Alan Aberdeen, Amelia Monks, Benjamin Moxley-Wyles, Emad Rakha, David R J Snead, Jens Rittscher, Clare Verrill

## Abstract

The use of immunohistochemistry in the reporting of prostate biopsies is an important adjunct when the diagnosis is not definite on haematoxylin and eosin (H&E) morphology alone. The process is however inherently inefficient with delays while waiting for pathologist review to make the request and duplicated effort reviewing a case more than once. In this study, we aimed to capture the workflow implications of immunohistochemistry requests and demonstrate a novel artificial intelligence tool to identify cases in which immunohistochemistry (IHC) is required and generate an automated request.

We conducted audits of the workflow for prostate biopsies in order to understand the potential implications of automated immunohistochemistry requesting and collected prospective cases to train a deep neural network algorithm to detect tissue regions that presented ambiguous morphology on whole slide images. These ambiguous foci were selected on the basis of the pathologist requesting immunohistochemistry to aid diagnosis. A gradient boosted trees classifier was then used to make a slide level prediction based on the outputs of the neural network prediction. The algorithm was trained on annotations of 219 immunohistochemistry-requested and 80 control images, and tested by 3-fold cross-validation. Validation was conducted on a separate validation dataset of 212 images.

Non IHC-requested cases were diagnosed in 17.9 minutes on average, while IHC-requested cases took 33.4 minutes over multiple reporting sessions. We estimated 11 minutes could be saved on average per case by automated IHC requesting, by removing duplication of effort. The tool attained 99% accuracy and 0.99 Area Under the Curve (AUC) on the test data. In the validation, the average agreement with pathologists was 0.81, with a mean AUC of 0.80.

We demonstrate the proof-of-principle that an AI tool making automated immunohistochemistry requests could create a significantly leaner workflow and result in pathologist time savings.

## INTRODUCTION

Prostate cancer (PCa) is the most malignancy in men worldwide [1, 2] and biopsies with suspected prostate adenocarcinoma contribute a significant proportion of the workload for surgical pathology centres. In many parts of the world, demands on pathology services are increasing and staff numbers are falling. In the UK, for example, a 2018 survey by the Royal College of Pathologists (RCPath) found that only 3% of surgical pathology departments have sufficient senior medical staffing and around a quarter of the workforce are moving towards retirement. The United Kingdom (UK) National Health Service (NHS) spends an estimated £27 million ($34 million) on locum and private services to cover this lack in service provision [3]. Over 60,000 prostate biopsies are carried out in the UK per year [4] and over one million in the United States of America [5]. With some prostate biopsy cases being allocated over an hour for reporting under proposed workload guidelines, this represents a significant workload burden [6].

The potential benefits of digital pathology (DP) and artificial intelligence (AI) have been well described [7] and it is clear that there could be much to gain from the introduction of workflow-based AI tools that do not affect established decision making in supporting prostate biopsy reporting. While a number of tools exist for automated prostate biopsy screening, detecting, and grading of tumours, some with regulatory clearance for diagnostic use [8-13], uptake of such tools remains relatively limited thus far. A digital pathology workflow is needed to enable AI and with increasing numbers of deployments in cellular pathology laboratories worldwide, the pace of uptake of AI should increase accordingly, although challenges still remain in their development and deployment [14].

In a digital workflow, AI can be used to assist pathologists as they screen prostate biopsy slides ultimately looking to confirm or exclude malignancy. An important adjunct to diagnosing Pca is the request of immunohistochemistry (IHC) for evaluating suspicious foci. An unmet need is the ability to triage slides, without waiting for a pathologist to review the case, identifying which cases cannot be signed out by review of Hematoxylin & Eosin (H&E) alone and need IHC. If such automated requesting could be achieved, the workflow could be significantly streamlined.

To diagnose PCa, pathologists search for a number of characteristic visual cues until enough features are found for confidently diagnosing malignancy. These features can be architectural and cytological. For instance, in acinar adenocarcinoma glands are infiltrative, often small in size compared to benign ones and crowded together. Cytologically, there are usually larger nuclei with one or more prominent nucleoli, often presenting perinucleolar clearing [15]. In a proportion of cases, the prostate epithelium presents some of the features described above, but not to an extent that can lead to a convincing cancer diagnosis by morphology alone, or the features are morphologically convincing, but due to small size of the lesion, IHC would be required for confirmation. As a number of benign mimics of PCa and conversely deceptively bland variants of PCa exist, IHC is often required [16, 17]. Some examples of such unclear morphology include Prostatic Intraepitheial Neoplasia (PIN) with smaller glands that could represent early invasion (sometimes known as ‘PIN Atyp’), areas of atrophy that are probably benign or areas that are suspicious of cancer, but too small for definitive diagnosis, i.e. atypical small acinar proliferation (ASAP).

The proportion of cases requiring IHC varies across institutions, ranging from 25% to 50% of total cases in some reports [18, 19]. Clinical guidelines recommend the use of basal cell marker IHC to detect loss of basal cells in the epithelial tissue. The absence of basal cells is the hallmark of malignant prostatic glands [18] and IHC is highly effective at reducing diagnostic uncertainty [19]. The main IHC markers recommended by the International Society of Urologic Pathology (ISUP) for routine diagnostic practice include CK5/6, 34BE12, P63 or a combination of basal markers and AMACR in a ‘‘cocktail’’ stain [20, 21]. Examples of prostate biopsies stained with H&E and CK5 are shown in Figure 1. Not every gland that lacks basal cells is malignant. For example in adenosis as few as 10% of the glands can show basal cells [22]. Thus the decision to request IHC is made on a focus or area-based level [23], and may be based on one or a number of foci of interest where the morphology is ill-defined.

**Figure 1:**
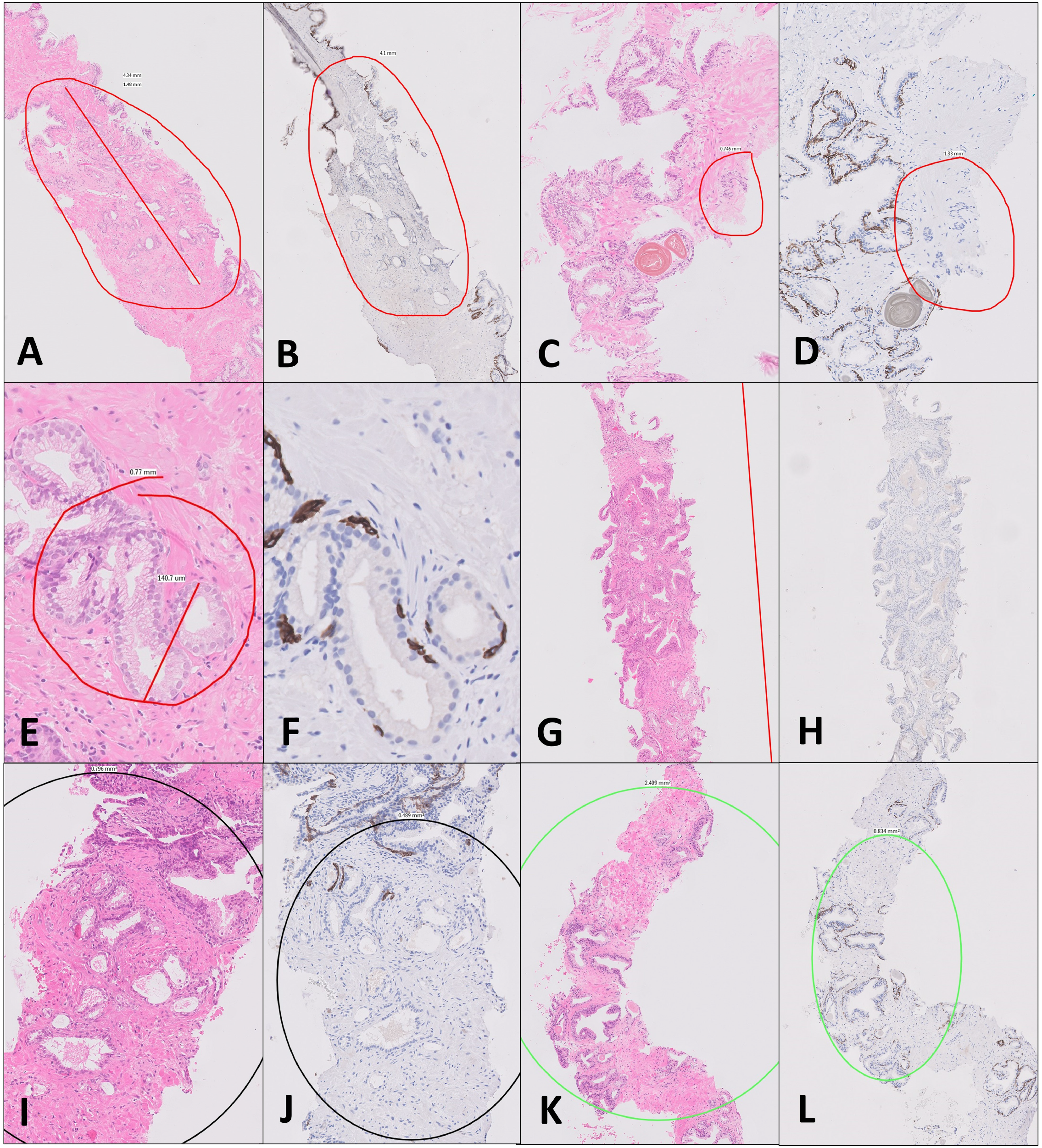
Examples of the classification of ambiguous prostate glands that prompt IHC ordering devised in this study (cf. Table 1). *Reason 1:* An H&E section showing a short length of glands (e.g., <1mm) that the pathologist is confident of calling cancer morphologically but wishes to confirm with IHC (**A**). This case was confirmed as cancer on CK5 staining (**B**). *Reason 2:* A small focus of glands that are only suspicious of are cancer (**C**). In this case, IHC confirmed cancer (**D**). *Reason 3:* Foci of glands with an unusual morphology and that the diagnosis would be uncertain (**E**). The CK5 in this case (**F**) demonstrated basal cell (brown) and the focus was deemed benign. *Reason 4:* A longer length of cancer (**G**) that is very well differentiated and needs to be confirmed cancer with IHC (**H**). *Reason 5:* Foci of glands that look atrophic but show atypia (**I**). In this example a few glands lack basal cells (**J**) but were considered benign as being admixed with this otherwise partially atrophic group. *Reason 6:* Small suspicious glands around PIN (**K**), in this example all were deemed benign (**L**). *Reasons 7 & 8:* not demonstrated here. These are cases used in this study and in some instances are annotated. Please ignore labels and arrows.

The request of IHC results in necessary delays to a case. Figure 2a illustrates the routine workflow for prostate biopsies common to most centres. The pathologist must first find time to review the morphology to make the decision and then put the case on hold while the IHC is performed. The time for IHC to be performed would vary across laboratories but is usually one to three days. When the pathologist reviews the case for further reporting sessions, there is inherent inefficiency and time wasted in refamiliarising oneself with the case.

**Table 1:** Reasons for ordering IHC staining on prostate core biopsy tissue slides with rates for each and resulting rates of malignancy. See also Figure 1 for examples.

| **Reason** | **Description** | **Number of foci identified** | **Number that were called cancer after IHC** |
| --- | --- | --- | --- |
| 1 | *A short length of cancer (e.g., <1mm) of any glands that show* | 187 | 168 |
| 2 | *Foci that are suspicious of cancer but only consist of a couple of* | 67 | 20 |
| 3 | *Unusual morphology glands that are difficult to classify* | 100 | 7 |
| 4 | *A longer length of cancer which has an unusual appearance (e.g.,* | 44 | 42 |
| 5 | *Foci that are atypical but probably benign (e.g., atrophy)* | 144 | 6 |
| 6 | *Small glands around PIN ? small foci of invasion or ASAP/PIN* | 64 | 6 |
| 7 | *Diffuse cells where the differential diagnosis lies between* | 32 | 0 |
| 8 | *Other – glands that do not easily fit into the above* | 3 | 0 |

**Figure 2:**
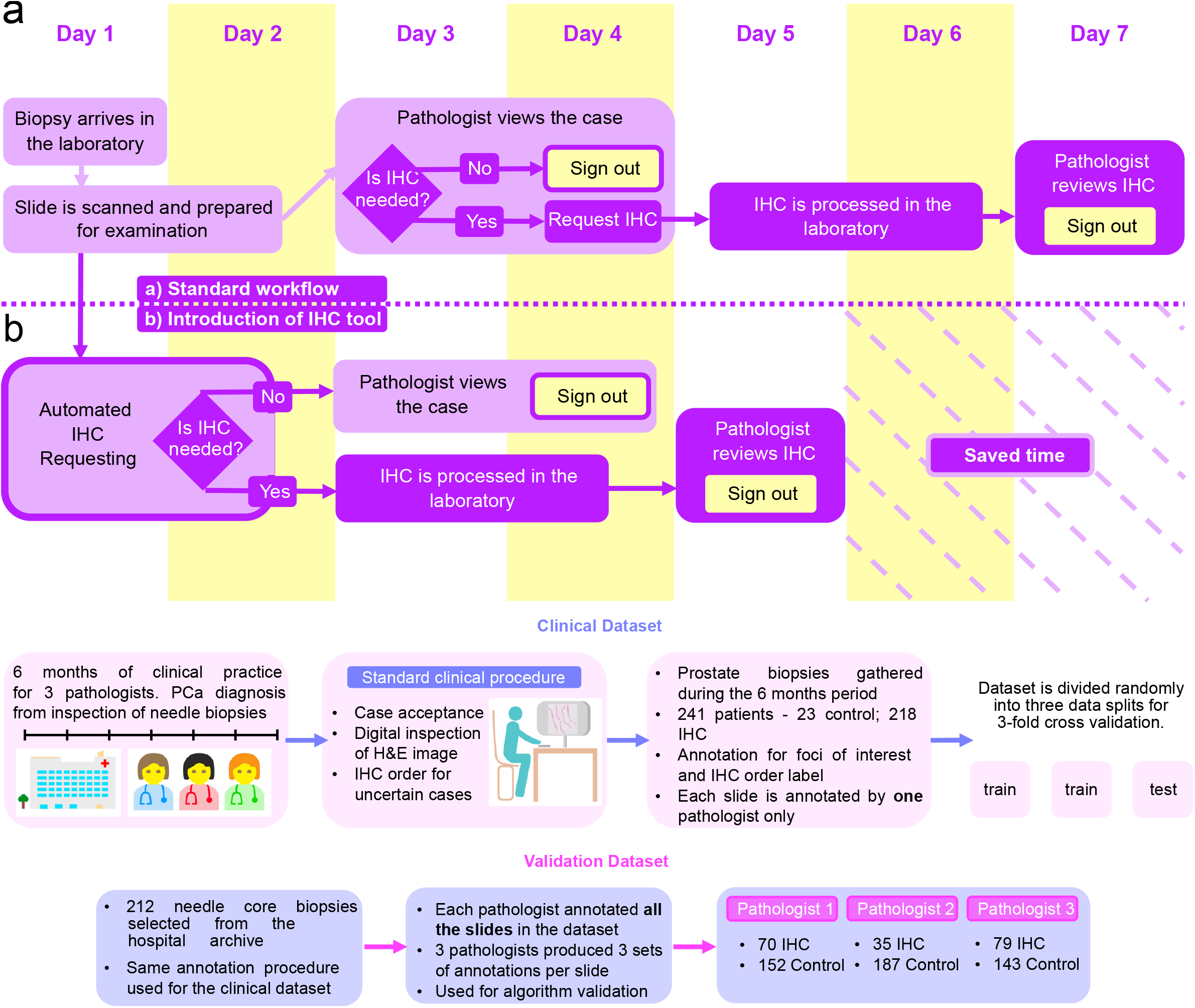
(a) Schema of typical workflow for diagnosis of prostate cancer from H&E-stained needle biopsies adopted by pathologists in the hospital. (b) Workflow after introduction of our tool for advance IHC requesting. The tool scans the H&E slide and requests IHC automatically, which is processed immediately after the H&E slide is scanned. The introduction of the IHC tool speeds up sign out of the case report. c) Datasets used for training, testing and validating the network. The dataset used for training and testing the network is a snapshot of standard clinical decisions made independently by pathologists. The validation dataset was annotated retrospectively and blindly to the archived data original labels. The procedure replicates the decision making that occurs in the hospital.

Our own experience confirms that IHC requesting increases turnaround time and reporting time for PCa and half of the delay due to IHC requests is incurred between the time the case is accepted by the pathologist and IHC is requested.

Here we design and validate an AI tool for automating the decision of requesting IHC for prostate biopsy cases. The study setting is one of the sites in the PathLAKE consortium, one of the UK Government’s AI Centres of Excellence. Several studies have demonstrated that novel computer vision algorithms based on high dimensional function optimisation called Deep Learning (DL) can extract visual features that encode clinically relevant morphological patterns from images, such as the Gleason score [8-13]. We build on these developments and demonstrate the training and validation of a DL system to individuate regions of prostate biopsy WSIs associated with diagnostic uncertainty. We then use the visual features extracted by the algorithm to train a Gradient Boosted Trees classifier for predicting whether IHC is required to diagnose a PCa case.

In this study, we demonstrate an AI tool which can trigger IHC requests from the H&E slides without the need for a pathologist to review the case first to make that decision. The pathologist then only needs to view the case once with all of the available stains and necessary delays to IHC requests are reduced. We describe the potential to expedite the prostate biopsy workflow, reducing inefficiency and ultimately reducing time to sign out, enabling results to patients and treating clinicians in a quicker time frame, as we propose in Figure 2b.

## MATERIALS AND METHODS

### Setting

The study was undertaken in a large academic teaching hospital in the UK (tertiary referral centre) with specialist urological pathology reporting and which processes 750-1100 prostate biopsies per year. The cellular pathology laboratory achieved the milestone of scanning 100% of the surgical histology workload in September 2020 with pathologists validated for full digital pathology reporting. Three specialist uropathologists (authors CV, LB, RTC) were involved in the development of the tool, two with greater than 10 years post-specialist registration experience, and one with 2 years post-specialist registration experience.

### Retrospective audits

In order to understand baseline rates of IHC request and potential workflow implications of the tool, all prostate biopsy cases were audited over a 12-month period, from August 2018 (before the introduction of digital pathology) to August 2019. The audit collected data on the case types (transrectal ultrasound guided biopsies or systematic transperineal biopsies), number of biopsies, turnaround times, extra work ordering, IHC requests and final diagnosis. To capture actual pathologist reporting times with and without IHC and necessary delays due to IHC request, a prospective audit of consecutive prostate biopsy cases reported by three specialist urological pathologists (CV, LB, RTC) during the period September 2019 to March 2020 was undertaken. For all cases, the date the case was received, the date the case was reviewed as H&E alone (reporting session 1), the date IHC was requested, the date of further reporting sessions where the case was reviewed with IHC (reporting session 2) and the date the case was signed out were recorded. Using stop-watches the following times were recorded: (1) length of time for initial slide review (H&E only) and make notes, (2) time to organise and make IHC request (3) if IHC requested, time to review case again with IHC, and (4) typing report.

### Modelling of potential time savings

In order to model potential time savings with upfront IHC ordering, we compared the mean turnaround times (date received to sign out) and pathologist reporting times (time for pathologists to examine and write report) for both IHC-requested and non IHC-requested cases. We assumed IHC would be performed shortly after H&Es were ready (e.g. process started within 3 hours) and not lead to any significant delays. Reporting time for IHC-requested cases was divided into two distinct sessions: during the first session, the pathologist examines the H&E slides and decides whether the case requires IHC. In the second session, the pathologist examines H&E and IHC slides together to make a diagnosis. The time savings that could be achieved by having IHC at the same time as H&E for reporting and thus removing duplication of effort is a complex one, including several factors in the decision making process. We calculated this for our laboratory in 2 ways: firstly, we assumed that the reporting time for cases with advance IHC requesting would be shortened to the reporting time for non IHC-requested cases. The second estimation method consisted in modelling the different tasks and factors impinging on reporting time individually. We compared the two estimates to approximate the benefits in reporting time.

### Prospective cohort curation

The study was conducted under the Pathology image data Lake for Analytics, Knowledge and Education (PathLAKE) research ethics committee approval (reference 19/SC/0363). We created a whole slide image (WSI) prostate biopsy training and testing set for the proposed IHC requesting tool from routine diagnostic cases. Prostate biopsies in which IHC for basal cell markers was requested during the period September 2019 to March 2020 were identified prospectively for study inclusion. Biopsies where IHC was not ordered before sign-out were excluded. Biopsies that could not be or had not been digitised were excluded. All biopsies were reported by one of three specialist uropathologists at our tertiary centre, using a mix of primary digital pathology reporting (Philips IntelliSite Pathologist Suite, Koninklijke Philips N.V, Amsterdam, Netherlands) and traditional light microscopy / glass slide reporting. All cases were digitally scanned Philips IntelliSite Ultra Fast Scanners (version 1.8.6614,20180906_R51, Koninklijke Philips N.V, Amsterdam, Netherlands).

### Classifying ambiguous foci

In order to understand the reasoning behind pathologist based IHC requests and identify categories to be modelled by machine learning, we devised a classification system of eight types of ambiguous prostate gland foci that would prompt IHC requests. These ‘reasons’ were based on the pathologists’ experience and were devised to include a representative range of the most common reasons for IHC ordering to confirm or exclude PCa. These are summarised in Table 1 with examples in Figure 1.

### Training Data

All cases which had had IHC requested by 3 reporting pathologists (CV, LB, RTC) as part of the diagnostic process during the period of the prospective study were included in training. Whole slide images (WSI) were de-identified using the Philips De-ID tool [version 1.1.5, Philips Digital Pathology Solutions Document DP-174226] and imported for annotation onto our in-house annotation platform, Annotation of Image Data by Assignments (AIDA) [24]. Each case was annotated by one of the pathologists only. All foci that prompted IHC ordering were included in the training dataset. In total, 299 WSIs for 241 patients were used to train the algorithm. Of these cases, 219 WSIs (187 patients) were non-selected consecutive cases that had prompted IHC ordering. The remaining 80 WSIs (54 patients) were selected from the previous (2019) clinical workload of 54 patients and designated as control cases. Pathologists annotated WSIs on the AIDA system. Pathologists drew around the focus (or foci) of interest (using a free hand digital drawing tool) that had prompted IHC ordering, and selected the reason for ordering IHC (up to 8 foci per case). Figure 2c summarises the collection and usage of WSI datasets. The control cases were reported as benign or malignant (50:50 split) and had had no IHC ordered at the time. We included these cases in the training/testing dataset in order to provide negative examples of benign and malignant cases for the algorithm.

### Algorithm development

We sought to develop an algorithm that could recognise tissue that is deemed ambiguous by pathologists and thus the case cannot be signed out by H&E morphology alone. We divided the histology data into a concise categorisation reflecting the decision process carried out in the clinic. In day-to-day practice, an IHC order is triggered by the presence of tissue with ambiguous morphology. Malignant tissue with very poor differentiation will take the organisation of higher Gleason patterns (e.g. amorphous sheets, cribriform glands), while benign tissue may mimic low Gleason patterns [3, 4]. As clearly benign, clearly malignant tissues are easy to distinguish by the pathologist, we group these tissues together as “certain” tissue. Intermediate differentiation levels are instead labelled as “ambiguous”. We trained a binary deep neural network (DNN) classifier to distinguish ambiguous from certain tissue. This corresponds to recognising all cases that cause sufficient uncertainty in the diagnostic procedure to require further information on the tissue, in the form of an IHC stain. The idea is illustrated in Figure 3a. We performed 3-fold cross validation in order to test the algorithm. We created 3 training splits of 200 slides by randomly sampling with replacement the 299 slides of our training dataset. The remaining 99 slides outside of each split were assigned as the test set. The ratio of control over ordered slides was fixed at 0.4 in each split.

**Figure 3:**
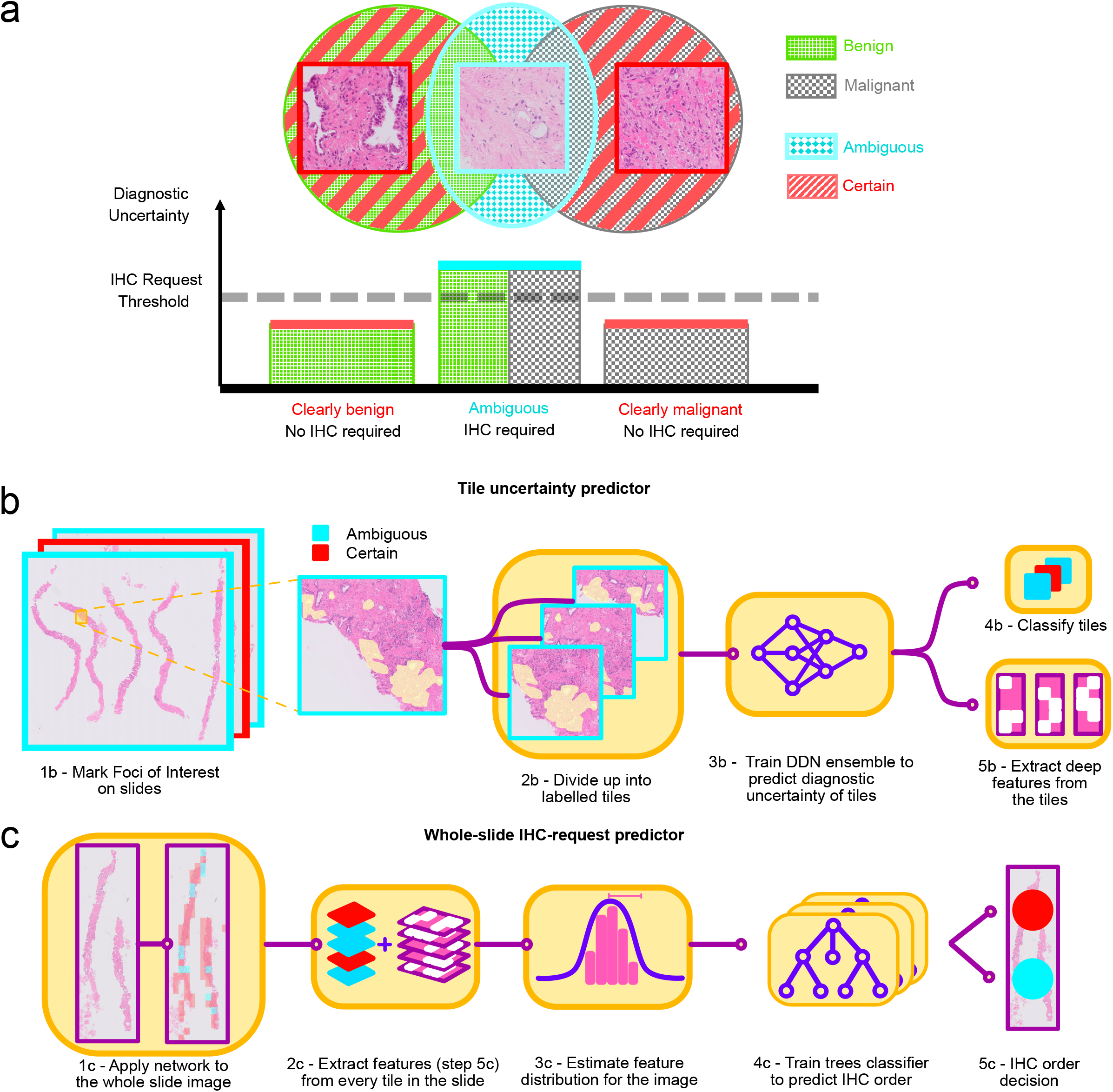
(a) Conceptual of IHC requesting. The Venn diagram illustrates the relationship between the benign vs malignant nature of tissue and the diagnostic need for IHC. Most tissue presents clearly benign (e.g. regularly-shaped glands) or clearly malignant morphology (e.g. amorphous tumour sheets). These cases cause relatively low diagnostic uncertainty, and IHC is not required to make a diagnosis (“certain” cases). A portion of benign and malignant cases presents morphology that cannot easily be placed in either class. Such “ambiguous” cases cause higher diagnostic uncertainty and require IHC for a diagnosis to be made. b, c) Decision making algorithm for ordering IHC for uncertain PCa cases. In part (b), a deep neural network ensemble is trained to recognise ambiguous epithelial morphology in tiles. The DNN also learns to compute representative features. In row (c) the morphology of whole slide images is characterised through the distribution extracted deep features, a gradient boosting classifier is trained to predict which cases require IHC for diagnosis.

Digital histology images can be corrupted by different types of artefacts. These include blur, debris and tissue folds, but also intensity irregularities in the backgrounds due to imprecision in the scanning process [25]. We trained a separate DNN in order to segmente tissue areas robustly.

1024×1024 tissue tiles at a resolution of 1.0 µm/px (10x) were extracted from annotated foci of interest within the tissue boundaries in the IHC-requested slides, and from benign and malignant regions in the control slides. An ensemble of three Residual Attention DNNs [26] was trained on each data split. The network ensemble was used to estimate the uncertainty of prediction on each tile, following the method described in [27]. The networks were trained for 200 epochs to convergence. Early stopping was not used. Instead, we relied on online space-domain and frequency-domain alterations of the training tiles, such as affine transforms and Gaussian noise, to augment the dataset and avoid overfitting. In order to evaluate the model, inference was performed on individual tiles on the foci, then the softmax class probabilities were averaged over all the tiles comprising the focus. The final label was assigned according to the class with the highest probability. The training procedure for the tile classifier is shown in Figure 3b.

Because there is no clear-cut criterion to determine whether the tissue morphology is atypical enough for the H&E stain to be diagnostically insufficient alone, the assessment contains a degree of uncertainty. This subjective component in the IHC order decision can lead to differences in annotations between pathologists. The conflicting tile labels can result in overfitting of the model over tissue features upon which pathologists disagree. Besides the uncertainty in tile labels, a pathologist can decide to order IHC for a patient because of multiple interesting tissue features present in different locations of the slide. Hence, an approach that considers each tile in the whole slide image separately is not sufficient to perform an accurate IHC order decision for the patient. Thus, a second step of the algorithm was designed to decide whether to order IHC that takes into account the tile-level features and prediction uncertainties aggregated over the whole slide. First, the contents of all tiles in the slide were compressed into a feature vector. The DNN was applied to all tissue tiles in every H&E image. Figure 4 shows examples of tile classification. Tile feature vectors were calculated from individual tiles through a similar approach to [28]. Distribution statistics (median, mean, variance, and kurtosis) were computed for features with an “ambiguous” label, and for tiles with a “certain” label. Furthermore, the prediction probability variance and loss variance for the slide were computed. The decision model consisted of a random-boosted-trees classifier, which was then trained on the slide feature vectors to predict whether IHC should be ordered for the patient case for IHC staining. Feature vectors were computed for each slide following the process detailed above, and the decision model was applied to all slides. The procedure is shown in Figure 3c.

**Figure 4:**
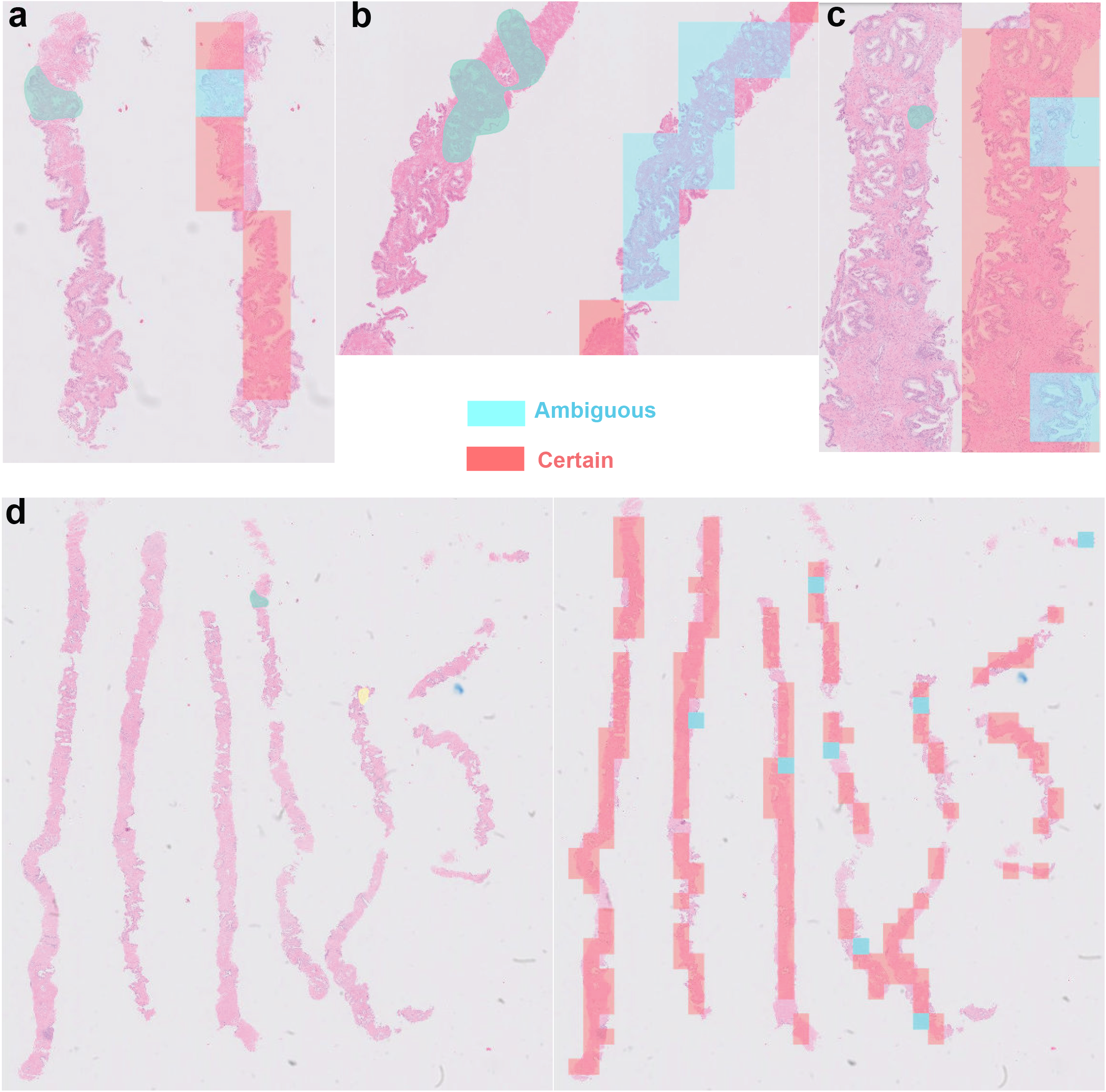
Example application of the tile classifier on the tissue areas of an unseen whole H&E slide (from the validation dataset). In (a-d), the results are compared to foci annotated by pathologists. Red denotes tiles classified as certain, while blue denotes tiles classified as ambiguous. (a), (b) and (c) show the overlap between pathologist annotation and the classifier output. The classifier marks as ambiguous regions that were identified by pathologists. (d) is an example of the tile classifier output for a WSI.

### Clinical Validation

A retrospective clinical validation of the algorithm on a fresh set of images was performed. For the validation, 100 new prostate biopsy cases were selected consecutively from the 2019 scanned slide archive. Cases that were not scanned were later excluded bringing the dataset to 91 cases. The total number of slides was 212. Cases were selected from early 2019 prior to the collection of annotated cases for training, in order to avoid dilution of the case mix with the removal of cases for training. In order to maximise the range of potential morphological appearances, while limiting the number of images requiring review, from each case one specimen / site was selected (specimen 1 of the case) and the H&E slides were used from one level of the tissue. Thus, one image was taken from each case and presented blindly to pathologists on AIDA. All three pathologists reviewed all images, thus generating 3 separate sets of validation annotations. Pathologists annotated the image to note if IHC ordering would be prompted or not for that image in their opinion, in the same manner as was performed for the training annotation set. The algorithm was then applied to all slides. The ResNet ensemble was used to classify all tissue tiles of the WSI. Features were computed as explained in the previous section and used to predict which cases need IHC requesting in the validation set. The algorithm decisions were then compared to pathologists’ decisions.

## RESULTS

### Turnaround and reporting times audits

The results of the retrospective audit enabled a comparison of necessary time costs incurred when IHC is requested for PCa cases. The mean turnaround times for IHC-requested cases was 7 days and 10 hours (95% CI: (7 days 2h, 7 days 16 h), n = 380), while the mean turnaround time for non IHC-requested cases was 4 days and 5 hours (95% CI: (4 days, 4 days 9 hours), n = 576). This included all types of prostate biopsy and the case mix was similar across both cohorts, as both IHC-requested and non IHC-requested cases had a median of 3 blocks. This indicates that the potential time saving by the introduction of this tool in our laboratory was 3 days and 5 hours on average (95% CI: (2 days 20 hours, 3 days 12 hours), n_1_ = 380, n_2_=576, Welch-Satterthwaite (WS) approximation [29]).

Non IHC-requested cases took an average of 17.9 minutes to be diagnosed (95% CI: (16.7 minutes, 19 minutes), n = 128), while the reporting time for cases where IHC was requested averaged at 33.4 minutes (95% CI: (30.7 minutes, 36.2 minutes), n = 133) over the course of two or more reporting sessions. The average time difference was therefore 15.5 minutes. The time savings that could be achieved by having IHC at the same time as H&E for reporting and thus removing duplication of effort are influenced by several factors inherent to the slide review and diagnostic decision processes. We calculated this for our laboratory in two ways: Firstly assuming automatic IHC ordering would reduce the reporting time for IHC-requiring cases to the same time taken to diagnose cases with no IHC request, we estimate a time saving of at least 12.6 minutes per case, by taking the lower end of confidence interval for the difference between mean IHC and non IHC requested reporting times, and 15.6 minutes on average (95% CI: (12.6 minutes, 18.5 minutes), n_1_ = 133, n_2_=128, WS approximation). Secondly, we assume a workflow where H&E review occurs during reporting session 1, and reporting session 2 consists of review of the IHC together with re-review of highlights of the H&E. In a slide viewing session, the pathologist screens the slides/images, spends time viewing difficult areas in more detail and makes a decision either to order IHC or make a firm diagnosis. In session 1 more time is spent on difficult areas and a decision is made to order IHC. In reporting session 2, the pathologist re-reviews the H&E, focusing on the areas of difficulty, reviews the new slides (IHC) and makes a decision. Thus in an IHC-requested case, the duplication points are re-reviewing the H&E to refamiliarise with the case in session 2 and making a further set of decisions than if the case were reported once with all of the necessary slides (as a decision is made in both sessions 1 and 2) and making this decision takes time.

We make an assumption that an additional step of decision making in an IHC case takes 1.5 minutes. During re-reviewing of H&E, the previously marked foci of ambiguous tissue are examined. Similarly, during first reviewing of IHC, corresponding foci are looked at to confirm staining status. We therefore assumed that the time taken to re-review H&E and for reviewing IHC are approximately equal. Thus, 7.5 minutes are spent re-reviewing the H&E and 7.5 minutes are spent reviewing the IHC. From our audit, IHC requesting took an average of 1 minute. We estimate therefore that 11 minutes can be saved by having IHC at the same time as H&E and the case only viewed once. This is likely to be a conservative estimate and does not take into account additional time picking up additional sets of slides from the lab, marking up additional slides etc.

### Annotation data

169 prospective cases were included in the study, from which 641 foci were identified across the three levels for each core that prompted IHC ordering. These foci were annotated for training the algorithm. Of these, Pathologist A annotated 32 foci, Pathologist B 284, and Pathologist C 325, which was in proportion to their clinical workloads.

The breakdown of the reasons for ordering IHC and the final diagnoses are given Table 1, with examples in Figure 1. The commonest reasons for IHC request were small foci of cancer needing confirmation (187 foci) and atypical foci that were probably benign (144 foci).

### Algorithm performance

The reliability of tile-level foci classification is reported in Table 2a. The ensemble attains excellent classification performance on the unseen test sets. Figure 4 compares example output of the tile classifier on unseen validation data with pathologists’ annotations. The model outputs a “certain” label for 90% of the test tiles. Hence, the models learned that only small regions of the needle biopsy contain ambiguous tissue morphology. This reflects standard diagnostic practice carried out by pathologists, where the need to order IHC is decided from small portions of the needle biopsy. While pathologists only individuate foci of ambiguity on slides where an IHC stain will be requested, the algorithm finds at least some ambiguous tiles in each slide of the dataset, with only 4 out of the 99 slides of the test set containing no ambiguous tiles. Thus the large number of ambiguous foci is likely to be due to morphological characteristics of tissue that were not present in the training data due to the potential range, and thus have never been seen by the model, which will produce spurious classifications for at least some tiles in most images. Most images are large, with a mean number of tiles in each image for the test dataset of 209, which increases the chance of encountering such different tissue appearance.

**Table 2:**
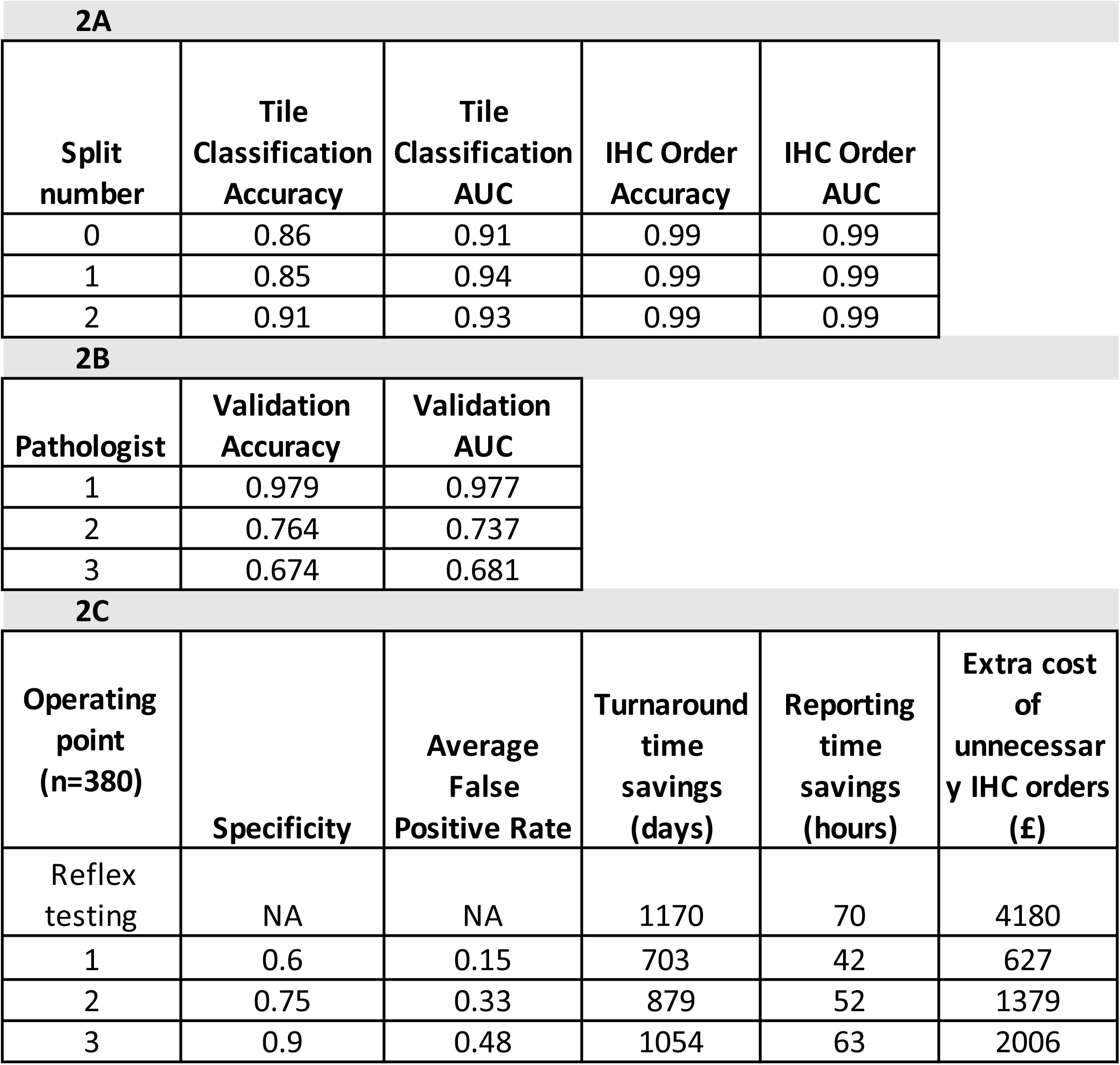
(a) Tile-classifier metrics for the task of classifying foci of interest on H&E slides, and slide-level classifier metrics for the IHC request prediction task, for the 3 cross validation splits of the dataset. (b) Slide-level classifier results for the task of IHC requesting evaluated on the validation set, for the 3 annotating pathologists. (c) Estimation of time savings and extra costs from introduction of the model (n=380 IHC requested cases). The false positive rate is averaged over the 3 pathologists. The minimum expected savings of 3 days 2 hours for turnaround time and 11 minutes for reporting times were used in the calculations. We assume an IHC order cost of £11 per slide.

These results highlight the need for a decision-making step that is robust to the presence of tissue regions with an “ambiguous” labelling in the image. The second step of the algorithm was designed to make a slide-level decision based on morphology and solve this issue. The IHC-ordering decision step performs well on the test set, as detailed in Table 2a (mean accuracy: 99%, mean AUC: 0.99).

The decision algorithm also predicts pathologists’ IHC order requirements on the validation dataset. The algorithm predicts very well the need for IHC staining according to all pathologists. Table 2b reports the agreement metrics for the IHC order decision between the model and each one of the three pathologists. In Figure 5, the receiver operating characteristic (ROC) curves for the model predictions vs pathologists’ annotations are reported. As a result of the disagreements in IHC ordering between pathologists, the model matches more closely the IHC ordering of pathologist 1, and performs the poorest when compared against pathologist 3’s opinion. The average agreement with pathologists is 0.81, with an average AUC of 0.80.

**Figure 5:**
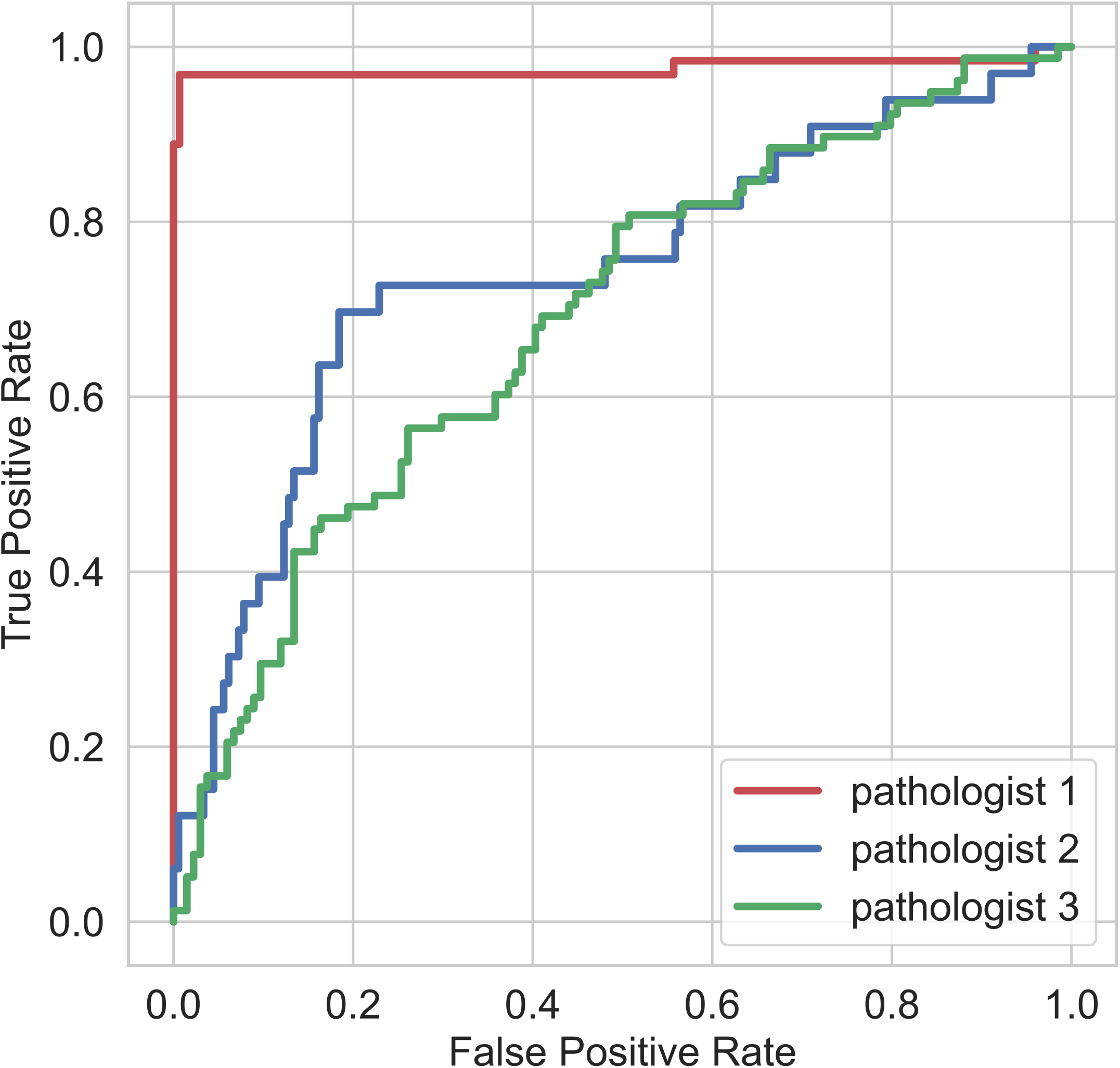
ROC curve for the tool vs the 3 pathologists (validation set). (d) Turnaround and reporting time savings, and extra incurred costs, for the 3 chosen operating points (at the 0.6, 0.7 and 0.9 values of specificity), for the 380 IHC-requested cases in the retrospective audit.

### Algorithm analysis

In order to understand what morphological details the model was sensitive to, we derived salience maps for foci from ordered slides and control slides with guided backpropagation [30] (see Figure 6a). The network examined the lumen structures inside prostate glands, the nuclei the gland is composed of, and the size of the epithelial cell bodies. Overall, these results highlight that the DNN is capable of recognising the salient features of epithelial structures. This is reflected in the feature vectors constructed from the DNN features and outputs from each slide, and used to train the slide-level IHC request classifier, whose projection onto the principal components is shown in Figure 6b. The vectors from the three datasets belong to the same point cloud in principal component space (Fig. 6c). Furthermore, the separation in feature space between IHC requested and non IHC-requested slides, albeit imperfect, suggests the vectors are representative of ambiguous/certain morphological features.

**Figure 6:**
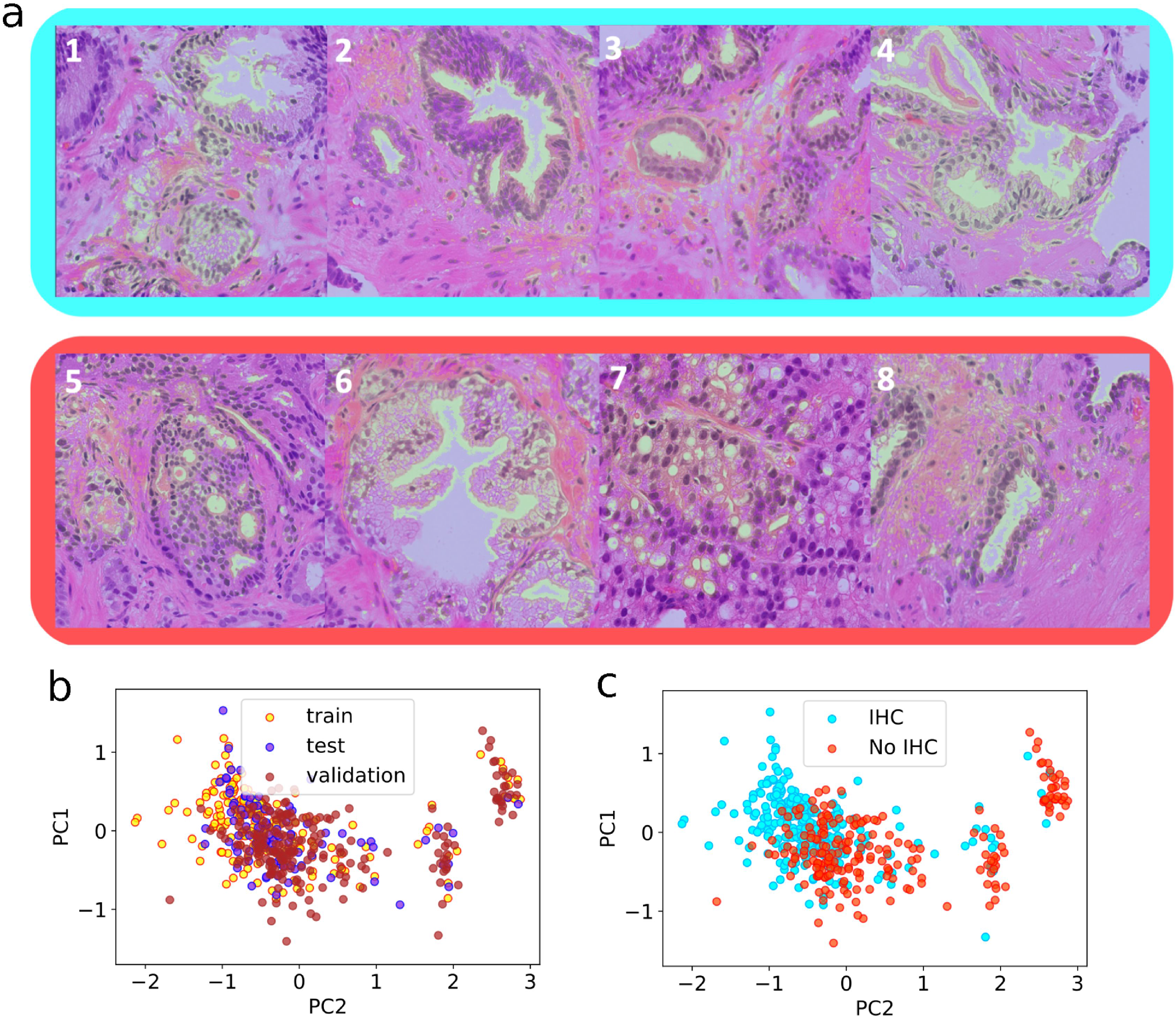
Results of two-step classification. The first step is the tile-level classifier. (a) shows salience maps (blue for ambiguous and red for certain cases), which highlight some morphological characteristics used by the tile-level classifier to distinguish between the two classes (obtained through guided-backpropagation [30]). The second step is the slide-level IHC request decision. (b,c) the manifold of slide feature vectors projected onto its principal components. Each data point is the feature vector summarising tissue content for one slide. Feature vectors are labelled according to the dataset of origin (b) and the IHC request status (c).

### Workload and time savings impact

We examine 3 potential operating points on the ROC curve, and the trade off between time savings and additional IHC requests that would be incurred if the tool was operated at those points. The points are marked on Figure 4 and correspond to a specificity of 0.6 (point 1), 0.75 (point 2) and 0.9 (point 3). Table 2c reports the time savings and additional incurred costs. Out of 974 retrospectively audited cases, IHC was requested for 380 cases, or 40% of total cases. We used the minimum predicted time turnaround time saving of 3 days and 2 hours and the minimum predicted reporting time saving of 11 minutes for the calculations, as discussed earlier. A higher specificity of the chosen operating point corresponded to larger savings in turnaround time and reporting time on one hand, and to larger extra costs due to overcalling on the other hand. Operating at a lower specificity yielded smaller predicted costs, but the predicted time savings were also reduced. The operating point with the highest specificity (0.9) provided a similar time saving to ordering IHC on all cases, but at half the cost of such reflex testing. Across 1000 sets of prostate biopsies needing IHC, we conservatively estimate the tool would save 165 pathologist hours.

## DISCUSSION

In this study, we evaluate the potential implications of automating the pre-requesting by AI of IHC in prostate biopsy cases that contain ill-defined epithelial morphology. Benign tissue and malignancies of the prostate present a large variety of morphological patterns, which pathologists must recognise by fitting the features to categories of recognized visual features and identifying distinctive characteristics of malignancy. Regions of tissue containing ambiguous morphology constitute a challenge to uropathologists, as a diagnosis cannot be made without IHC and even with IHC, the tissue may remain in an ambiguous category such as ASAP.

We developed a novel composition of a deep learning system to detect ambiguous versus certain morphology in needle core biopsies and a boosted random forest to predict which cases require IHC using the tissue content representations encoded by the deep learning system. The deep learning system was trained on pathologists’ annotations of ambiguous tissue regions, and it successfully recognised image foci contributing to diagnostic uncertainty on H&E WSIs. The gradient boosted trees classifier was used to mimic the decision-making process of pathologists and predict which slides require IHC. This second step was needed because most cases have at least some ambiguous areas flagged reflecting the variety of morphological appearances that can be seen in the prostate. The slide-based classifier is needed to translate information about ambiguous tissue content in the whole slide into a decision about which cases are sufficiently ambiguous to require IHC.

The IHC-request decision step correctly predicted the IHC request decisions on the test set obtained through 3-fold cross validation. The algorithm was also validated on the independent validation dataset, where it satisfactorily matched the IHC request decisions of 3 different pathologists. The good classification results obtained on the validation dataset point to good generalisation properties of the network. The agreement rate between pathologists in the validation set was between 61% and 64%. This is consistent with reports of interobserver variability in other diagnostic tasks with a subjective component, such as Gleason reporting (consistently reported to be around 60% [31, 32]). In order to simulate over and under-ordering scenarios in a potential real-life deployment of the tool, we set 3 arbitrary points on the ROC at specificity levels of 60, 75 and 90%, resulting in false positive rates (over-ordering of IHC) in 15, 33 and 48% of slides respectively. Taking the most conservative level of specificity, 40% of cases that needed IHC might be missed by the tool and it might over order on 15% of slides.

We took the approach to deliberately target diagnostically ambiguous cases rather than basing the tool on a tumour detection tool to better reflect the inherent complexity of the decision to request IHC. Although a tool based on tumour detection could be set to identify small foci of morphologically convincing tumour or longer lengths of unusual but certain cancers (reasons 1 and 4) and request IHC for confirmation, these scenarios accounted for 36% of cases which was far fewer than the ambiguous areas where the diagnosis was not certain which accounted for 59% cases (reasons 2, 3, 5, 6, 8). Some of these cases may fall through the net in a tumour-finding approach being classified as benign with no IHC required. Although most areas had a gland-based morphology, reminiscent of Gleason patterns 3 and 4, rather than diffuse sheets of cells which may represent pattern 5, 5% of the annotated foci fell into this latter category. Regardless, the tool performed well when applied onto either pattern. The training also lacked other non-adenocarcinoma diagnoses of the prostate, such as urothelial carcinoma, potential neuroendocrine carcinoma or soft tissue lesions due to absent or infrequent training examples, which may be addressed in future iterations of the tool.

Requesting IHC involves additional tissue staining and allocating extra pathologist time for case re-examination, which increases organisational complexity. In this study we showed that prostate biopsies requiring IHC take double the time to be reported by pathologists, and twice as many days to be reported. Some of this is process related, but also it is recognised that cases requiring IHC are inherently more complex. A few characteristics of the IHC workflow contributed significantly to higher time costs. Firstly, we found that the time between case reception and the IHC request date is an average of 2 days, which is redundant time whereby cases are awaiting review. Our tool moves the decision to the point at which the H&E is created in the lab. The time to perform the IHC and the inherent complex nature of these cases does not change with this tool. Rather, we have shown that 11 minutes per case can potentially be saved by advance IHC ordering, which we believe to be a conservative estimate. This is achieved by reducing the inefficiencies of 2 reporting sessions in re-reviewing slides and duplicating decision making, i.e. the time-consuming decision of ordering IHC before the final diagnostic decision can be made. Processing the IHC slides in a contiguous time slot to the H&E slides provides a leaner workflow for the lab as the case is only booked out to the pathologist once. Our approach involves targeted advance IHC requesting: request on every case should not be adopted in practice because the costs involved in staining extra tissue for every case outweighs the benefits [33].

Like all similar tests there is inevitably a trade off in this model, with some degree of over or under calling. We envisage that the tool would be used adaptively and that centres would be able to select an acceptable threshold for ordering IHC based on institutional preference. We would need to explore with regulatory bodies how to achieve the setting of different performance points within the appropriate regulatory framework. This would likely involve submitting the validation data for a number of set thresholds to support the intended purpose. Laboratories would then be required to verify performance to the satisfaction of their governance team and external laboratory accreditation bodies.

The roadmap to introduction of AI into cellular pathology is complex, which has thus far limited uptake [14]. In this study, we describe a proof-of-concept algorithm for IHC request by AI that effectively takes a workflow step away from the pathologist but does not directly affect the diagnosis. In particular in the case of under-calling by the tool, i.e. a missed IHC request, IHC can nevertheless be requested by the pathologist after visual assessment of the H&E slide. Hence this is a relatively low risk task, which might serve as a good entry point for the use of AI.

There is an inherent component of subjectivity in the IHC requesting task, and thus a ground truth is difficult to define. A tool trained by a group of pathologists from one centre might not entirely represent practice in another centre and the decision to request may be affected by a number of other non-morphological factors – level of fatigue, pathologist and institutional experience, degree of specialisation, psychological and personality factors. The next iteration of the tool will need careful design to capture multi-centre training and validation and establishment of the ground truth by a panel of pathologists. For this we will leverage our fortunate position within the PathLAKE digital pathology consortium (one of the UK Government’s AI Centres of Excellence) [34]. In outlining likely workflow benefits and economic impacts, we acknowledge the limitations of our dataset and that the workflow we describe here may be slightly different in other institutions. A fully developed tool will ultimately require prospective validation in a real-time health care setting, with a wider evaluation of what time savings are deliverable in practice.

One aspect of practice that could be considered as a weakness of this AI tool, is that in some difficult cases the thinking/reflecting time afforded by waiting for IHC is helpful. Of course, the tool does not stop the pathologist from walking away from a case for a while to get perspective, if clinically appropriate. In the many cases where the IHC is for confirmation of what is already a confident diagnosis on H&E this should not be an issue.

In the future, this work could be expanded for application to other prostate settings (such as transurethral resections). There is also potential to apply the tool to other tissue types (e.g. breast or lung biopsies), or to develop a generic tool for automated IHC or molecular requests.

In summary, we designed and evaluated a tool for advance IHC requesting with the potential to reduce diagnostic times for PCa in the clinic. Our algorithm emulates the decision-making of pathologists and robustly estimates when IHC staining is required to diagnose a prostate biopsy. Unlike previous work that focuses on predicting the presence of cancer, we focus on automating a routine clinical task at the core of accurate PCa diagnosis. We believe tools that help pathologists carry out their daily tasks and improve clinical workflow will provide significant benefits to healthcare institutions and expedite the adoption of digital pathology in cancer clinics worldwide.

## Data Availability

The datasets generated during and/or analysed during the current study are not publicly available due to the terms of the PathLAKE Consortium Agreement but are available via the corresponding author on reasonable request.

## ACKNOWLEDGEMENTS

The authors would like to thank Innovate UK, UK Research and Innovation (UKRI), the National Institute for Health Research (NIHR) Oxford Biomedical Research Centre (BRC), and the Department of Cellular Pathology, Oxford University Hospitals NHS Foundation Trust (OUH). We acknowledge the contribution to this study made by the Oxford Centre for Histopathology Research and the Oxford Radcliffe Biobank which is supported by the NIHR Biomedical Research Centre.

This study was conducted in the setting of Oxford University Hospitals NHS cellular pathology laboratory which scans 100% of the surgical histology workload and enabled this study. The authors would like to thank all of the staff who contributed to this significant achievement which was a team effort involving the biomedical scientist, secretarial and pathology staff among others.

Manuel Salto-Tellez and Jacqueline A James are Principal Investigators in PathLAKE at Queens’s University Belfast and Nasir Rajpoot is a Principle Investigator in PathLAKE at the University of Warwick -all were involved in generating the PathLAKE programme, including funding.

CV and LB are part funded by the National Institute for Health Research (NIHR) Oxford Biomedical Research Centre (BRC). The views expressed are those of the author(s) and not necessarily those of the NHS, the NIHR or the Department of Health.

Views expressed here are those of the authors and not necessarily those of the PathLAKE Consortium members, the NHS, OUH, Innovate UK or UKRI.

## ETHICS APPROVAL AND CONSENT

The study was conducted under the PathLAKE research ethics committee approval (South Central Oxford C Research Ethics Committee (UK), reference 19/SC/0363) and consent to data research was checked. Patients are not identifiable from the material. The research was performed in accordance with the Declaration of Helsinki.

AUTHOR CONTRIBUTIONS

(authors listed in same order as title page list)

Conceptualisation: AC, RC, LB, JR, CV; Data Curation: AC, RC, LB, NKA, SM, MH, AM, BMW; Writing (original draft): AC, RC; Methodology, software: AC; Formal analysis: AC, RC; Audit planning: RC, CV; Annotation: RC, LB, CV; Funding Acquisition: ER, DRJS, JR, CV; Writing (editing): AC, RC, CV; Writing (reviewing): AC, RC, KS, SM, LB, JR, CV; Supervision: JR, CV; Visualization software: AA

